# Using the illness-death model to estimate age- and sex-standardized incidence rates of diabetes in Mexico from 2003 to 2015

**DOI:** 10.1101/2023.12.27.23300547

**Authors:** Omar Silverman-Retana, Ralph Brinks, Annika Hoyer, Daniel R. Witte, Thaddäus Tönnies

## Abstract

**Objectives:** To estimate the age-specific and age-standardized incidence rate of diabetes for men and women in Mexico between 2003 and 2015, and to assess the relative change in incidence of diabetes between 2003 and 2015.

**Methods:** We use a partial differential equation describing the illness-death model to estimate the incidence rate (IR) of diabetes for the years 2003, 2009 and 2015 based on prevalence data from National Health Surveys conducted in Mexico, the mortality rate of the Mexican general population and plausible input values for age-specific mortality rate ratios associated with diabetes.

**Results:** The age-standardized IR of diabetes per 1000 person years (pryr) was similar among men (IRm) and women (IRw) in the year 2003 (IRm 6.1 vs. IRw 6.5 1000/pryr), 2009 (IRm: 7.0 vs. IRw: 8.4 1000/pryr), and in 2015 (IRm 8.0 vs. IRw 10.6 1000/pryr).

**Conclusions:** The incidence rate of diabetes in Mexico between the years 2003 and 2015 remained stable. However, rates were markedly higher among women in the age group 40-49 and 50-59 in the year 2015 compared with rates in 2003.

## Introduction

Mexico ranked 7 in the top 10 countries with the number of adults aged 20-79 years living with diabetes in 2021 (1). The country has the highest age-adjusted prevalence within North America and the Caribbean Region (16.9%, representing 14.1 million citizens). Furthermore, it has been estimated that 6.7 million of Mexican citizens lived with undiagnosed diabetes in 2021. Mexico ranks 8 in the top 10 leading countries with the highest total health expenditure (USD 19.9 billion) due to diabetes in adults aged 20-79 years (1).

The epidemiology of diabetes in Mexico has been extensively documented in terms of the disease prevalence (2–9). Since early in the 1990s, reports from National Health Surveys (carried out every six years since the year 2000) showed a steady growth in the prevalence of diagnosed diabetes from 4.6% in 1993 to 11.1% in 2020 (3,4). Moreover, the geographical variation of type 2 diabetes prevalence shifted from being observed predominantly in the Northern states to the Southern states over a three decade period, with a higher annual average growth among younger age groups compared with the growth observed in older age groups (6). Finally, the number of Mexican citizens living with diabetes has been projected to reach between 15 and 25 million individuals by 2050 (2).

In contrast, few studies have investigated the incidence rate of diabetes among the Mexican adult population (2,9). Studies showed that the age-specific incidence rate of diabetes exponentially increased in both sexes during the period from 1970 to 2010, and that these rates doubled every ten years (2). Findings from The Mexico City Diabetes Study (MCDS) reported an incidence rate of type 2 diabetes of 14.4, and 13.7 per 1000 person-years (pryr) for men, and women respectively in the age group of 35 to 64 years old (9). Knowledge about incidence of diabetes is relevant to understand the dynamics of diabetes at the population level and to identify target populations for possible prevention interventions. Therefore, we aimed to estimate the age- and sex-specific as well as the age-standardized incidence rate of diabetes in Mexico in 2003, 2009 and 2015, and further assess the relative change in incidence of diabetes between 2003 and 2015.

## Methods

The estimation of the incidence rate of diabetes without individual follow-up data is based on the mathematical relation between the age-specific prevalence of diabetes, the mortality rate of the general population, and the mortality rate ratio (*MRR*) associated with diabetes. Specifically, we used a partial differential equation which governs the illness-death model of chronic diseases (10). This approach has been used previously for estimating the incidence rate of diabetes, the incidence of diabetes complications, and the incidence rate of other chronic diseases (11–14).

### Estimating the incidence rate of diabetes

Brinks and Landwehr developed a partial differential equation (10), which describes the temporal change in prevalence with respect to age and calendar time. Solving this equation for the incidence rate *i* yields:

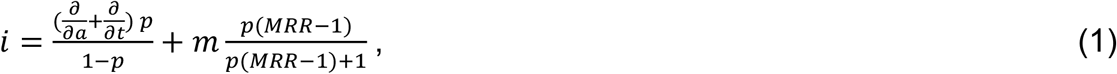

where *p* represents the prevalence of diabetes, 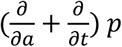 the temporal change in prevalence with respect to age *a* and calendar time *t, m* the mortality rate of the general population and *MRR* the mortality rate ratio. Equation (1) illustrates that the incidence rate *i* can be calculated by plugging in estimates for *p*, its temporal change 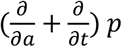, *m* and *MRR* on the right hand side of the equation. To estimate the temporal change in *p*, cross-sectional data on age-specific prevalence for at least two years are required. As described below, age-specific prevalence for the years 2000, 2006, 2012 and 2018 was available for this study. We used these data to estimate the incidence rate midway between these cross-sectional studies, i.e. the years 2003, 2009 and 2015. Due to small sample size in age groups ≥80 years, we restricted the estimation of the incidence rate to age groups 20-79 years. The mortality rate of the general population (*m*) for the years 2003, 2009 and 2015 was based on publicly available data from official population vital statistics retrieved from the National Statistics and Geography Bureau (“*INEGI*-Instituto Nacional de Estadística y Geografía” from its acronym in Spanish- https://www.inegi.org.mx/datosabiertos/). Unfortunately, no nationally representative data are available for MRR. As in previous analyses (13), we considered a plausible range of age-specific MRR ranging between 7.5-8.5 at age 30 years, and 2.5-3.0 at age 80 years. Between ages 30 and 80 years we assumed a log-linear relation between age and MRR as observed in previous studies (15,16). The ranges of age-specific MRRs are in line with one study including participants from Mexico City (17), and studies from other countries (15,16).

To account for the uncertainty in the input data we calculated 95% confidence intervals (95% CI) based on a resampling approach (18). We repeated the estimation of the incidence with Equation (1) 2,000 times. For each repetition, we drew samples from the distribution of the input prevalence, which was estimated with logistic regression as described in section 2.2. MRR values at age 30 and 80 years were drawn from uniform distributions in the range of plausible values described above. We report the median, 2.5^th^ and 97.5^th^ percentiles of the resulting distribution of incidence estimates as the point estimate and 95% CI.

### Estimation of input prevalence

We used publicly available data from the Mexican National Health and Nutrition Survey (ENSANUT for its acronym in Spanish, data available here: https://ensanut.insp.mx/) to estimate the self-reported age- and sex-specific prevalence of diabetes for the years 2000, 2006, 2012, and 2018. In brief, ENSANUT is a probabilistic survey of Mexican households that covers a broad range of health and nutritional indicators. The complex survey design allows making statistical inference of the health component at national and subnational levels. The detailed study design of each wave of ENSANUT has been documented elsewhere (19). We estimated the prevalence in each survey year separately with logistic regression models that accounted for the complex survey design using the survey weights provided by ENSANUT. To account for the non-linear relationship between age, calendar time and diabetes prevalence we used a natural cubic spline with five equally spaced knots for age. The logit-transformed prevalence from the logistic regression models for the different survey years were combined into one data set and used as the dependent variable in a linear regression model with sex and a B-spline with 3 knots for age as independent variables. We also included a linear term for the survey year to account for temporal changes in prevalence. To allow for age- and sex-specific prevalence as well as age- and sex-specific trends in prevalence we included a three-way interaction between age, survey year and sex. Modeling the logit-transformed prevalence from the different survey years in one regression model was necessary to appropriately account for the complex survey design and at the same time obtain the smoothed input data for the partial differential Equation (1). All analyses were carried out with the statistical software R, version 4.1.3 (20).

## Results

The ENSANUT data included 178,514 participants of which 14,965 reported to have been diagnosed with diabetes (Table 1).

**Table 1.** Description of the ENSANUT Mexico study population for the years 2000, 2006, 2012 and 2018.

| **Characteristic** | **Total**<br>**(N = 178,514)** | **No Diabetes**<br>**(N = 163,549)** | **Diabetes**<br>**(N = 14,965)** |
| --- | --- | --- | --- |
| Age (yrs) | 43 (16) | 42 (16) | 57 (13) |
| Sex (women) | 89,175 (50%) | 80,972 (50%) | 8,203 (55%) |
| Survey year |  |  |  |
| 2000 | 44,078 (25%) | 41,123 (25%) | 2,955 (20%) |
| 2006 | 45,140 (25%) | 42,175 (26%) | 2,965 (20%) |
| 2012 | 46,277 (26%) | 41,787 (26%) | 4,490 (30%) |
| 2018 | 43,019 (24%) | 38,464 (24%) | 4,555 (30%) |

Table shows mean(SD) or numbers(%)

Figure 1 shows the age-specific prevalence estimated with these data using logit-transformed prevalence estimates. The prevalence from Figure 1 was used as input for equation (1) alongside input data for the mortality rate of the general population and the MRR to estimate the age-specific incidence rate shown in Figure 2.

**Figure 1.**
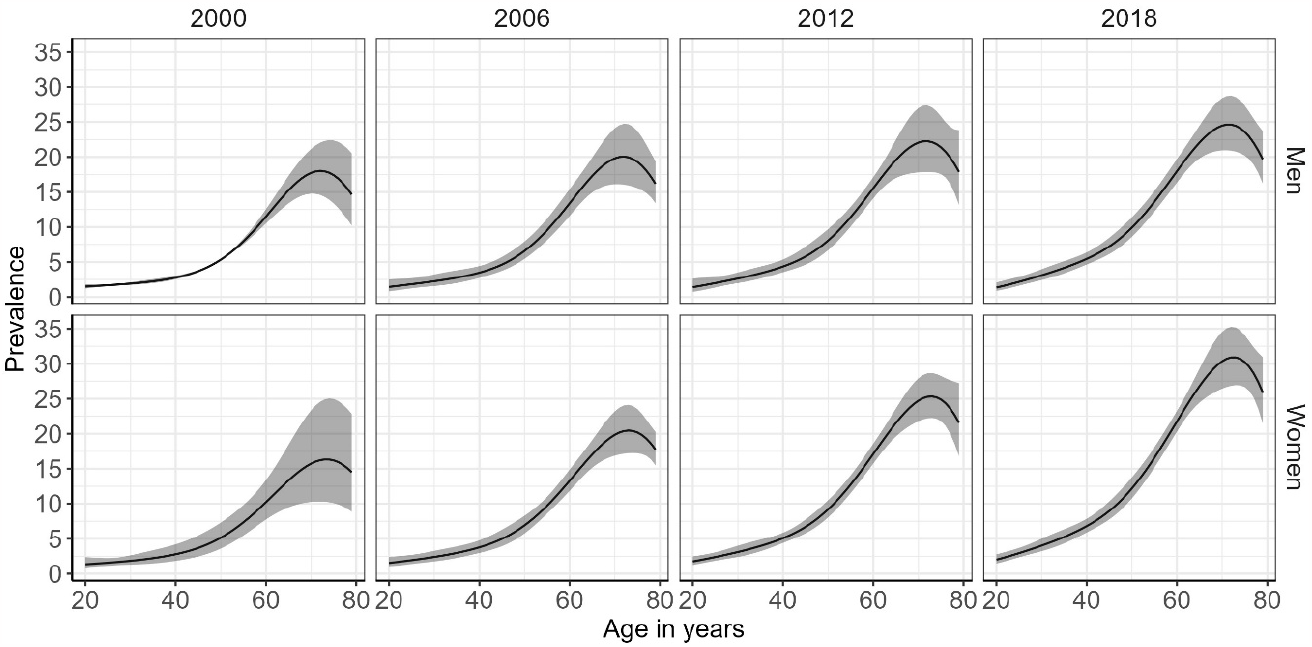
Age and sex-specific prevalence of diabetes in Mexico in the years 2000, 2006, 2012 and 2018 Prevalence was estimated with regression models after logit-transform (solid lines) with 95% CI (shaded areas) accounting for the complex survey design and survey weights.

**Figure 2.**
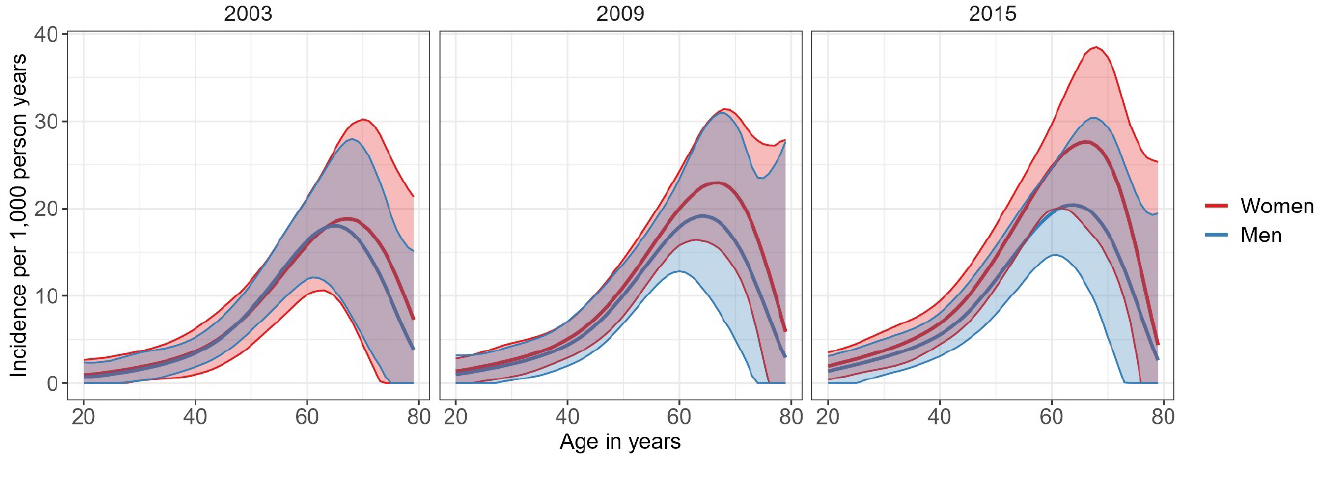
Age- and sex-specific incidence rates of diabetes in Mexico in the years 2003, 2009 and 2015 Shaded areas represent 95% CI

The results indicate an increase in incidence rates of diabetes among men and women between 2003 and 2015 in almost all age groups >40yrs. The incidence rate of diabetes showed a steep increase until the age of ∼65 years, where it peaked, and gradually decreased.

Table 2 shows the temporal change in age-standardized incidence rates, and for 10-year age groups. The age-standardized incidence rate increased by a factor of 1.61 (95% CI: 0.89-4.03) among women, and by 1.32 (95% CI: 0.66-2.66) among men between the years 2003 and 2015. In general, the results are rather imprecise, which is indicated by the wide confidence intervals.

**Table 2.** Age-standardized-and age-specific incidence rates of diabetes (95% CI) per 1,000 pryr for men and women in Mexico in the years 2003, 2009 and 2015, and the temporal change in incidence rates.

| Year | Incidence per 1,000 PY (95% CI) |  |  | Change in Incidence (95% CI) |  |
| --- | --- | --- | --- | --- | --- |
|  | 2003 | 2009 | 2015 | IRR<br>(2009 vs. 2003) | IRR<br>(2015 vs. 2003) |
| **Men** |  |  |  |  |  |
| **Age-standardized** | 6.1 (3.5-9.2) | 7.0 (3.7-11.1) | 8.0 (4.5-11.8) | 1.15 (0.54-2.36) | 1.32 (0.66-2.66) |
| **Age groups** |  |  |  |  |  |
| 20-29 | 1.1 (0.1-2.6) | 1.6 (0-3.4) | 2.1 (0.0-4.0) | 1.37 (0.00-11.91) | 1.89 (0.01-16.7) |
| 30-39 | 2.3 (0.9-4.1) | 3.1 (0.9-5.3) | 4.0 (1.9-6.2) | 1.32 (0.34-3.84) | 1.71 (0.68-5.14) |
| 40-49 | 5.4 (3.4-7.7) | 6.7 (3.8-10.1) | 8.1 (5.0-11.4) | 1.23 (0.63-2.28) | 1.49 (0.83-2.65) |
| 50-59 | 12.5 (9.2-16.3) | 14.2 (10.6-18) | 16.1 (11.7-20.4) | 1.14 (0.77-1.68) | 1.28 (0.86-1.9) |
| 60-69 | 18.0 (10.6-26.3) | 19.1 (10.2-29.7) | 20.3 (12.5-29.3) | 1.06 (0.52-2.12) | 1.13 (0.61-2.2) |
| 70-79 | 9.7 (1.5-19.3) | 9.3 (0.0-23.5) | 9.6 (0.0-21.6) | 0.94 (0.00-6.81) | 0.97 (0.00-7.91) |
| **Women** |  |  |  |  |  |
| **Age-standardized** | 6.5 (2.8-10.2) | 8.4 (5.2-11.7) | 10.6 (7.1-14.5) | 1.28 (0.66-3.14) | 1.61 (0.89-4.03) |
| **Age groups** |  |  |  |  |  |
| 20-29 | 1.4 (0.2-3.1) | 2.0 (0.2-3.6) | 2.8 (1.1-4.5) | 1.35 (0.14-8.89) | 1.94 (0.61-15.02) |
| 30-39 | 2.6 (0.6-4.6) | 3.6 (1.6-5.4) | 5.0 (2.6-7.3) | 1.38 (0.51-5.58) | 1.91 (0.85-7.90) |
| 40-49 | 5.5 (2.3-8.7) | 7.5 (4.7-10.2) | 9.9 (6.9-13.1) | 1.35 (0.69-3.37) | 1.78 (1.01-4.59) |
| 50-59 | 12.0 (7.3-16.3) | 15.6 (12.1-19) | 19.7 (16.0-23.7) | 1.29 (0.87-2.15) | 1.63 (1.14-2.77) |
| 60-69 | 18.5 (9.9-26.8) | 22.7 (16.2-29.9) | 27.5 (18.7-36.6) | 1.23 (0.73-2.41) | 1.47 (0.86-2.99) |
| 70-79 | 14.0 (2.0-25.8) | 14.7 (2.7-27.5) | 15.6 (3.7-28.1) | 1.06 (0.18-7.34) | 1.10 (0.27-8.66) |

IRR: incidence rate ratio. Incidence in 10-year age groups was estimated using the midpoint of the age groups (e.g. age 55 years for age group 50-59 years)

## Discussion

Based on the mathematical relationship between prevalence, incidence and mortality (10), our study contributes to the current knowledge on diabetes epidemiology in Mexico by presenting age- and sex-specific, and age-standardized incidence rates of diabetes between 2003 and 2015, and the relative change in incidence in this period.

Overall, the relative change in the age-standardized incidence rate of diabetes among men and women was not different between the years 2003-2009, and 2003-2015. Yet, we found a 78% and 63% increase in the incidence of diabetes among women in the age groups 40-49 and 50-59 years respectively, between the years 2003 and 2015. Recent reports on the global incidence of diabetes showed stable or decreasing trends on incidence rates of diabetes across different populations (1).

Our results are estimated incidence rates, modeled based on sequentially observed prevalences. Conceptually, increases in observed diabetes prevalence are due to a mix of longer survival of diabetes patients, demographic changes in the population (larger proportions in high-risk age groups), improved diagnosis (emptying the pool of undiagnosed cases) and a true increase in age-specific incidence (21). A Danish register-based study has tried to disentangle the relative contributions of some of these sources, and estimated that 9% of the prevalence changes were due to decreasing mortality rates, 27% due to imbalance between incidence and mortality and 22% due to increasing incidence (22). Our mathematical model accounts for changes in mortality and demographic structure, while our incidence estimates still include both the contribution of improved diagnosis as well as of a true age-specific increase in incidence. A true age-specific increase in incidence rates is likely due to a higher prevalence of the major risk factors for diabetes at an earlier age. Mexico has a very high prevalence of overweight and obesity [19], particularly affecting women. The risk of type 2 diabetes in overweight and obese Mexicans aged 50 years and older is double and triple the risk of Mexican individuals with normal weight, respectively [20]. Yet, we can only speculate that unhealthy weight could be a strong driver in this specific women’s age group, as our model only took age and sex into account.

Our estimates are smaller than those previously reported for the Mexican population. The Mexico City Diabetes Study reported an incidence rate of type 2 diabetes of 14.4/1000 pryr among men and 13.7/1000 pryr among women in the year 2008. These differences may be explained by the fact that the MCDS recruited participants from a low-income, urban population aged between 25-64 years at recruitment (9); while we used data from a national representative survey covering a broader age range (20 to 80 years). Meza et al. used the self-reported date of diagnosis to estimate the age-specific incidence rate of diabetes (2). This approach is subject to selection bias, as people living without diabetes are more likely to survive until the survey date than people living with diabetes. This “immortal time bias” is introduced as people did not die between the diagnosis date and the survey date, and hence were “immortal” during that period when they were included. One of the main strengths of our study is that we used a method to estimate the age-specific diabetes incidence rate that is not prone to immortal time bias, since only cross-sectional data on an aggregated level is required as input data. Furthermore, we used survey data in our model that has national representativeness; therefore, our results can be extrapolated to the Mexican adult population. As a drawback, we relied on self-reported diabetes status and therefore a potential misclassification of the outcome could bias the result towards the null. This misclassification might bias our estimates only if men reported (or participated in the survey) differently from women, if the case, it would be difficult to ascertain the direction of the bias. Furthermore, it is not possible to distinguish between type 1 and type 2 diabetes, which is particularly relevant for younger age groups.

Our results might be partly driven by trends in undiagnosed diabetes. The most recent Global Diabetes Atlas (2021) estimates that in Mexico 47.5% of cases are undiagnosed, the 6th country with highest undiagnosed proportion, while earlier editions in 2019 and 2017 put this estimate at 38.6% and 37.4% respectively (1,23,24). However, it should be noted that the sources used for the Diabetes Atlas have been updated with every edition and may have employed different methodologies. Globally it is unclear whether the diagnosed proportion is increasing, stable or decreasing. Analysis of sequential NHANES data suggests that in the USA the undiagnosed proportion has fallen from 19.3% around 1990 to 9.5% around 2020 (based on a combined assessment of fasting plasma glucose and glycated hemoglobin-HbA1c), and hence emptying of the pool of undiagnosed cases has contributed to the rise in prevalence of diagnosed diabetes in these years (25). Other analyses of the same NHANES data using different definitions report a higher undiagnosed proportion, but also a fall over time. Previous studies from Mexico suggest, like the Global Diabetes Atlas, that the undiagnosed proportion in Mexico is increasing (3,4).

The number-needed-to-screen (NNTS)-a prevalence-based measure for “goodness-of-detection” (26), was 45 in 1993, 55 in 2000, 13 in 2006, 20 in 2012, 22 in 2016, 14 in 2018, and 19 in 2020. This means that in the year 1993, the reciprocal of the average number of persons without a diagnosis a physician must see to meet one undiagnosed case was 45, whereas in 2020 this number was 20. In fact, the smaller NNTS observed in recent years point towards a deterioration of diabetes case-finding strategies over time. Another possibility could be that incidence is happening in younger age groups and that the same strategy is yielding diminishing results. It is therefore unlikely that the trends in diabetes incidence we observed were primarily due to improvements in detection of undiagnosed diabetes. Regarding the MRR as input data, we had to rely on data from the National Danish Diabetes Register due to the lack of representative data for Mexico. However, based on an argument from Breslow & Day, the MRR is stable across different populations (27).

In conclusion, we showed similar incidence rates of diabetes among the Mexican adult population over the period from 2003 to 2015. Yet, a substantial increase was observed among Mexican women aged 40-69 years.

## Data Availability

We used publicly available data. ENSANUT data can be accessed here: https://ensanut.insp.mx/. Vital statistics data and census data can be accessed here: https://www.inegi.org.mx/datosabiertos/

https://www.inegi.org.mx/datosabiertos/

## Acknowledgements

OS-R is supported by a research grant from the Danish Diabetes Academy, which is funded by the Novo Nordisk Foundation, grant number NNF17SA0031406. OS-R received support provided by the Steno Diabetes Center Aarhus (SDCA), which is partially funded by an unrestricted donation from the Novo Nordisk Foundation.

## Contributors

OS-R and TT made substantial contributions to the conception and design of the study. OS-R was in charge of the acquisition of the data, and TT performed the statistical analysis. RB and AH contributed to the analysis and interpretation of the data. OS-R and TT drafted the manuscript. RB, AH, and DRW critically revised the manuscript for important intellectual content. OS-R, TT, RB, AH and DRW approved the final version of the manuscript. TT is the guarantor of this work.

## Ethics approval

This is a secondary data analysis of publicly available data. ENSANUT surveys were approved by the committees of Ethics and Research of the National Institute of Public Health of Mexico.

